# Artificial intelligence-generated digital Romberg test for peripheral neuropathy monitoring

**DOI:** 10.64898/2026.05.12.26353015

**Authors:** Clara Tejada-Illa, Ariadna Pi-Cervera, Jordi Pegueroles, Mireia Claramunt-Molet, Ainhoa Heras-Delgado, Jesus Gascón-Fontal, Sebastian Idelsohn-Zielonka, Mari Rico, Núria Vidal-Fernández, Lorena Martín-Aguilar, Marta Caballero-Ávila, Cinta Lleixà, Roger Collet-Vidiella, Josué Moreno, Tania Mederer-Fernandez, Laura Llansó, Álvaro Carbayo, Ana Vesperinas, Luis Querol, Elba Pascual-Goñi

**Affiliations:** Institut de Recerca Sant Pau (IR SANT PAU), Barcelona, Spain; Neuromuscular Diseases Unit, Hospital de la Santa Creu i Sant Pau, Universitat Autònoma de Barcelona, Spain; Centro para la Investigación Biomédica en Red en Enfermedades Raras (CIBERER), Madrid, Spain; Ephion Health, Barcelona, Spain; Unit of Digital Health, Eurecat, Centre Tecnològic de Catalunya, Barcelona, Spain

**Keywords:** Digital biomarker, peripheral neuropathy, balance impairment, wearable sensors, Romberg

## Abstract

**Background and Objectives:** Patients with peripheral neuropathies (PN) commonly exhibit balance impairment. In clinical practice, balance is typically assessed using the Romberg’s test and ataxia scales, which rely on examiner interpretation, while objective biomarkers for quantifying balance remain lacking. Wearable sensors are valuable tools for objectively quantifying gait abnormalities in PN patients and may capture clinically meaningful changes over time. By integrating these parameters, artificial intelligence (AI) can assist in generating a digital score that enables easy, objective, and reproducible monitoring of patients’ postural balance. This study aims to generate and assess an AI-generated digital Romberg’s test to quantify balance impairments in a cohort of PN patients.

**Methods:** PN patients were assessed in a longitudinal study using a wearable system composed of inertial sensors placed on the trunk and plantar pressure sensors integrated in insoles. Patients performed the Romberg’s test under both eyes-open and eyes-closed conditions and were classified according to ataxia severity (mild, moderate, or severe) following the score obtained in item 1 of MICARS and SARA scales.

**Results:** We included 97 patients with PN (including autoimmune and hereditary polyneuropathies), and 117 healthy controls (HC). Significant differences in trunk sway and center of pressure (COP) were observed between groups, particularly with eyes closed. Using wearable sensor parameters, we developed an AI digital Romberg’s test, which correlated with clinician-rated Romberg’s test performance and distinguished patients with and without ataxia (AUC=0.632) and across different PN pathologies. Longitudinally, digital Romberg’s test and iRODS showed concordant trajectories. Also, changes ≥25% in the score were associated with clinical changes in ataxia severity measured by an increase in MICARS-SARA score (+1.42 points), whereas improvement was associated with a decrease (−0.20 points) in the scale.

**Discussion:** This study demonstrates that wearable sensors are useful to detect and quantify balance impairment. The AI-generated Romberg’s test is an objective and reproducible tool for postural balance assessment, with robust discriminatory performance across clinical ataxia severity in PN. Score’s longitudinal changes aligned with clinical severity, supporting its potential for monitoring disease progression and treatment response. Its strong association with balance measures reinforces its role as a quantitative biomarker of postural control in ataxia patients.

## Introduction

Balance and gait impairments due to sensory ataxia are common in patients with peripheral neuropathy (PN) and result in elevated risk of falls.^1,2^ Sensory ataxia is a neurological syndrome caused by impaired sensory feedback from peripheral nerves^3^ and characterized by loss of balance and coordination, particularly in the absence of visual input.^4^ A range of different balance and gait measures are used in clinical practice and clinical trials to assess treatments’ effectiveness. Balance is qualitatively evaluated in the neurological exam with Romberg’s test,^5,6^ where loss of postural control with eyes closed indicates proprioceptive deficits in the lower limbs.^7,8^ This assessment has some limitations^9^ and is inherently subjective^10^ as it relies on neurologist’s interpretation. Objective biomarkers capable of quantifying balance and serving as reliable outcome measures remain lacking.

In a previous study, we demonstrated that wearable sensors are valuable for monitoring gait abnormalities in patients with PN. Digital biomechanical features (DBF) capture differences between ataxic and steppage gait patterns, grade the severity of gait impairment and detect clinically relevant longitudinal changes.^11^ This system has also been successfully evaluated for gait analysis in other conditions, including Pompe disease.^12^ Balance has been previously assessed in patients with other neurological diseases using inertial measurement units (IMUs)^13,14^ and pressure platforms in patients with diabetic peripheral neuropathy,^15^ multiple sclerosis^16,17^ and other autoimmune^18,19^ and hereditary neuropathies.^20^ However, the potential of multiparametric sensors to detect balance impairments in a heterogeneous cohort of PN patients has not been explored.

This study aimed to assess and quantify balance deficits using wearable sensors. We used AI to generate a digital Romberg’s test that quantifies balance impairment with a simple metric and assesses the correlations between the digital Romberg’s test and conventional clinical scales in patients with PN, while also exploring its ability to capture meaningful changes over time.

## Materials and methods

### Patients

Participants were recruited among patients routinely followed at our center. Inclusion was based on a predefined clinical criteria, which required a confirmed diagnosis of PN established through clinical examination, electrophysiological studies, and genetic or immunological testing. Eligible participants included individuals with inflammatory neuropathies, such as chronic inflammatory demyelinating polyneuropathy (CIDP), monoclonal gammopathy patients of undetermined significance associated with IgM (IgM-MGUS), multifocal motor neuropathy (MMN) or chronic ataxic neuropathy, ophthalmoplegia, immunoglobulin M [IgM] paraprotein (CANOMAD), other autoimmune nodopathies and hereditary neuropathies including Charcot-Marie-Tooth (CMT). In addition, participants were required to exhibit clinical evidence of balance or postural instability, defined by abnormal findings in routine neurological examination.

### Protocol of the study

Patients were monitored over two years, with follow-up visits scheduled every six months for stable individuals and every three months for those experiencing a relapse or a treatment change. Patients who could not complete two years of follow-up were also included in this study. The study protocol is summarized in Figure 1.

**Figure 1.**
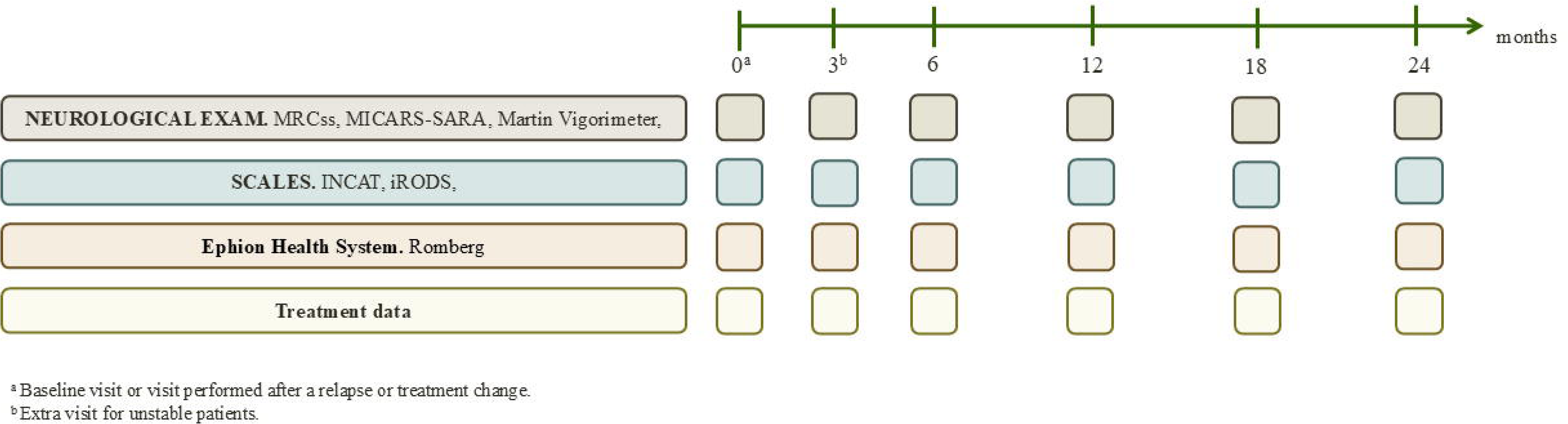
Protocol of the study. Monitoring visits performed every 6 months during 2 years for those patients who remained stable and for those who presented a relapse or a treatment change, a 3-month visit (3*) was performed after that date. MRCss: Medical Research Council sum score, INCAT: Inflammatory Neuropathy Cause and Treatment, iRODS: Inflammatory Rasch-built Overall Disability Scale, MICARS-SA.RA: Modified Internacional Cooperative Ataxia Ra6ng Scale and Scale for the Assessment and Rating of Ataxia.

During visits, the evaluating neurologists performed a neurological evaluation including the following scales: Medical Research Council sum score (MRCss), Inflammatory Neuropathy Cause and Treatment (INCAT), Modified International Cooperative Ataxia Rating Scale (MICARS), Scale for the Assessment and Rating of Ataxia (SARA), and grip strength testing via a Martin vigorimeter. Patients filled in the Inflammatory Rasch-built Overall Disability Scale (iRODS) questionnaire. Finally, Romberg’s test was performed while participants were wearing biomechanical sensors. Raw data from all wearable sensors were recorded via a dedicated smartphone application and automatically uploaded to a secure, cloud-based research database for storage and subsequent analysis. Ataxia was assessed using a face/content validity preselected list of items originating from MICARS^21^ and SARA,^22^ as implemented in the IMAGiNE protocol,^23^ a longitudinal study evaluating anti-MAG neuropathy patients. Each test performed by patients was classified by the evaluating neurologist according to ataxia severity (mild, moderate or severe) based on item 1 related to gait of the MICARS-SARA scale and prespecified clinical criteria (Supplementary Table 1). Postural control was further characterized using items 5 and 6 of the MICARS scale, which evaluate postural stability in the standing position through the Romberg’s test under different sensory conditions: eyes open (item 5) and eyes closed (item 6). Definitions and scoring criteria are provided in Supplementary Table 2.

### Wearable technology

We used the Ephion Mobility system (Ephion Health, Barcelona, Spain),^24^ which supports multiple sensor configurations. For this study, we relied on the information provided by three sensors: a trunk-mounted inertial sensor (Movesense, Vantaa, Finland)^25^ and two instrumented insoles (Moticon, Munich, Germany).^26^ Each device integrates an inertial measurement unit, while the insoles also include plantar pressure sensors. All sensors were synchronized via Ephion Mobility software on a smartphone. Participants wore the system while performing the Romberg test, enabling continuous capture of gait and balance variables. Although the full protocol included additional sensors^11^ (previously published), only balance-related biomechanical features were analyzed for this study (summarized in Supplementary Table 3).

### Data processing

Balance-related signals were obtained from two complementary parameters provided by the 3 sensors: (1) pressure changes under the feet, measured by insoles and expressed as the center of pressure (COP), and (2) trunk motion, estimated from an inertial sensor placed on the chest. While COP reflects how body weight is distributed and adjusted over the support surface, trunk displacement captures the overall movement of the body’s center during standing. For each signal we calculated an ellipse containing 95% of the recorded points.^27,28^ This ellipse represents the area within which the patient’s body oscillates during the Romberg test, providing an intuitive measure of postural stability: larger ellipses indicate greater sway and, potentially, poorer balance control.

When the corresponding ellipse showed abnormally large dispersion or irregular shapes, which are typically indicative of measurement artifacts (e.g., sensor misplacement or noise), data from that sensor were excluded from the analysis, ensuring that subsequent analyses were based only on reliable data.

From the remaining signals, we extracted features that characterize different aspects of postural control. For trunk motion, features included dominant frequency of sway (reflecting how fast the patient oscillates), the amplitude of movement (how much the patient moves), and variability (how consistent or irregular the movement is). Together, these measures provide information on both the magnitude and regularity of postural movements.^29^ For both insole and trunk data, ellipses were used to derive descriptive metrics such as area, amplitude, and shape. These parameters provide a concise and clinically interpretable summary of each patient’s balance behavior during the test.

To select the most representative features related to both open and closed eyes, before the statistical analysis, features with low variance and those containing more than 30% of missing values were excluded. After this data cleaning step, a total of 48 features remained from an initial set of 75. These cleaned features were then used in subsequent analyses.

### Digital Romberg’s test analysis

The digital Romberg’s test was computed using an in-house machine-learning pipeline. The dataset was split at the subject level, using 70% of subjects for model training and 30% for validation, ensuring that no individual contributed data to both sets. Within the training set, K-nearest neighbours imputation was applied to handle missing values, and feature selection was performed using an Elastic Net–regularized model, which combines L1 and L2 penalties to enforce sparsity and coefficient shrinkage. This procedure identified a stable subset of features strongly associated with MICARS–SARA 5 and 6.

Independent eXtreme Gradient Boosting (XGBoost) regressors were trained on the selected features for each clinical item, with hyperparameters optimized using cross-validation folds within the training set, again ensuring subject-level separation between folds. Model performance was evaluated on the held-out 30% of subjects using mean absolute error (MAE), root mean square error (RMSE), normalized root mean square error (nRMSE), and correlation with observed clinical scores, providing unbiased estimates of generalization.

The resulting digital Romberg’s test ranges from 0 to 100, where higher values indicate movement patterns consistent with the control population, and lower values reflect increased variability or postural instability.

### Statistical analysis

To examine the relationship between features and clinical assessments, patients were stratified according to their scores on MICARS-SARA item 1, and MICARS items 5 and 6. A Linear Mixed Effects (LME) model with patient-specific random intercept was first used to assess the differences between the control group and the PN group in both eyes opened and eyes closed separately, and those who were able to perform the test without such compensatory behaviors were included. Only features that showed significant differences between patients and controls were retained and subsequently used as input for the second set of LME models, used to determine which features were related to MICARS and SARA items 1, 5, and 6 and the total sum score of the clinical scale. This was also assessed for the two conditions. A significance threshold of p < 0.05 was applied, and results were corrected for multiple comparisons using the false discovery rate (FDR) method to ensure robust control of Type I error.

To assess the discriminative ability and clinical validity, group comparisons between controls and patients, as well as among different severity levels, were performed using Kruskal–Wallis tests with post-hoc pairwise comparisons. Associations between the score and clinical measures were examined using Spearman’s rank correlation coefficients. Receiver operating characteristic (ROC) curve was used to determine discriminative performance of the score between ataxic and non-ataxic patients. Finally, Spearman’s rank correlation coefficients were calculated between all clinical scales, patient-reported outcomes (PROMS) and the computed balance score.

To evaluate the longitudinal behavior of the proposed digital Romberg’s test and its relationship with clinical progression, the analysis was performed on all available pairs of visits for each patient of the test set. For each pair of time points, only pairs showing a relative change ≥25% were retained for further analysis. These changes were classified as improvement or worsening depending on the direction of the score variation. For each selected pair, the corresponding clinical values (total MICARS-SARA score) at the initial and final time points were extracted. Descriptive analyses were performed separately for improvement and worsening groups by comparing baseline and follow-up clinical values. Mean values and their 95% confidence intervals were calculated at each time point to summarize the distribution of clinical scores. A complementary longitudinal analysis was conducted using the iRODS centile as the clinical anchor. In this case, all available visit pairs were retained regardless of the magnitude of relative change in the digital score. For each patient, the overall trajectory of iRODS centile scores over time was classified into three groups according to the MCID threshold of 8 points^30^: worsened (net change ≤ −8), improved (net change ≥ 8), or stable (|net change| < 8). Linear mixed-effects models (LME) were then fitted separately for each trajectory group to characterize the evolution of both the iRODS centile and the digital Romberg score over elapsed time.

### Data availability

The datasets generated and/or analyzed during the current study are available from the corresponding author upon reasonable request.

## Results

### Baseline characteristics of the study population

Ninety-seven patients with peripheral neuropathies and 54 healthy controls were included in the study. The mean age of participants was 60.1 ± 12.6 years, and 61 patients (62.9%) were male. Patients presented different types of neuropathies, including 43 with CIDP (44.3%), 5 CANOMAD (5.2%), 22 with IgM-MGUS (22.7%), 9 with MMN (9.3%), 10 with CMT (10.3%), and 8 with other autoimmune nodopathies (8.2%). Detailed baseline characteristics are summarized in Supplementary Table 4.

### Wearable sensor-derived static balance features

This analysis evaluated differences in wearable sensor–derived static balance during Romberg test performance under eyes-open and eyes-closed conditions across ataxia severity groups. Results for COP and trunk sway according to ataxia severity are shown in Figure 2. The margins of the insoles show the distribution of points across different areas of the insole, allowing the visualization of how patients’ postural control differs from controls. In eyes open condition (Figure 2A), COP distributions and ellipses corresponding to trunk sway increased according to ataxia severity (for mild and moderate groups), but differences remain relatively compact except for an increase in sway area in trunk movement in one patient from the moderate ataxia group. With eyes closed (Figure 2B), differences were more evident across severity groups (for mild and moderate groups), showing larger ellipse areas in COP and trunk variables reflecting a marked increase in sway amplitude and variability. In the severe group, trunk sway was reduced, with smaller ellipse areas compared to other severity groups. Two complementary analyses were conducted to characterize wearable sensor-derived balance features. First, biomechanical parameters were compared between PN patients and healthy controls to identify features associated with balance impairment. Second, the relationship between these features and ataxia severity was evaluated. LME were applied separately for the eyes-open and eyes-closed conditions, accounting for repeated measures within subjects. The resulting regression coefficients (β) and corresponding p-values are summarized in Supplementary Tables 5 and 6. Of the 47 features analyzed, 10 features derived from COP, and 12 from trunk sway showed significant associations after correction for multiple comparisons, particularly in the eyes-closed condition, indicating that increased sway amplitude, velocity, and variability are associated with greater postural instability. These significant features were used to analyze the associations across ataxia severity, with 7 features related to COP and 10 related to trunk showing significant differences according to ataxia severity.

**Figure 2.**
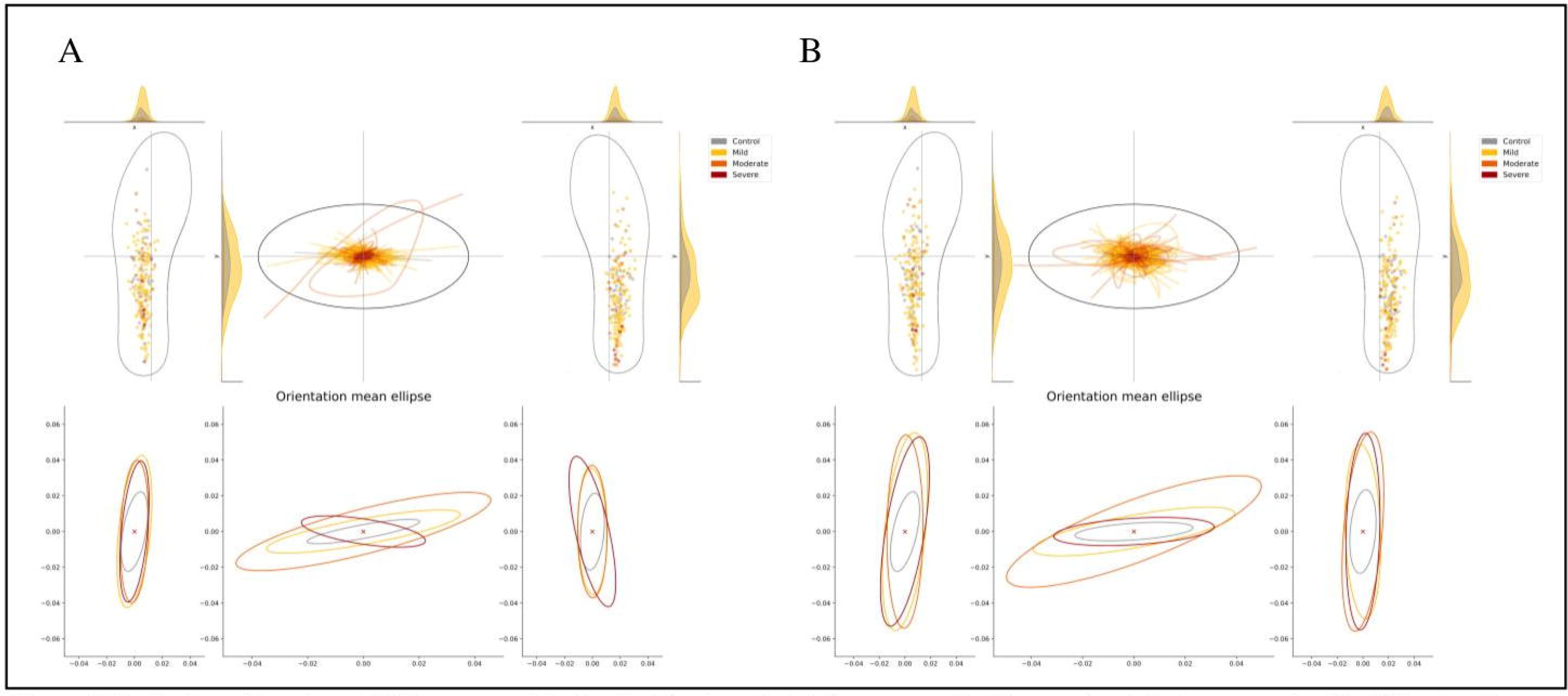
Evaluation of trunk and COP movement during the Romberg test. (A) eyes opened, (B) eyes closed. Patients were classified following the score obtained in item 1 of MICARS and SARA scale. At the top of each figure, there are displayed the insoles which represent the ellipse centers as ‘dots.’ In gray, the area where the controls maintain the center of their ellipses and, in color, the subject’s central position during the functional tests. The upper central graph represents the movement generated by the trunk during the test, enabling visualization of the trunk’s position over time. At the bottom of each figure, the left and right graphs correspond to the ellipses from the insoles. The lower central graph corresponds to the ellipses from the trunk.

### Digital Romberg’s test

To integrate all relevant features in a single metric, a balance score, the digital Romberg’s test, was generated. The digital Romberg’s test performance was evaluated by examining its relationships with the total MICARS and SARA scores as well as specific subscale items, including item 1 (general ataxia), item 5 (Romberg eyes open), and item 6 (Romberg eyes closed). Scores closer to 1 indicated patterns similar to those of the control group, whereas lower scores reflected increased variability and postural instability. Significant differences were observed between the healthy control and PN, with patients exhibiting substantially lower scores (Figure 3A). Additionally, the digital Romberg’s test significantly discriminated amongst ataxia severity levels (Figure 3B). The ability of the digital Romberg’s test to discriminate between patients with and without ataxia was also evaluated. For this purpose, the presence of ataxia as determined by physician’s assessments was used for each patient and session. The digital Romberg’s test achieved an AUC of 0.632 (Figure 3C), indicating modest discriminative performance between ataxic and non-ataxic patients. For the comparison between controls and patients, ROC analysis demonstrated good discriminative performance, achieving an AUC of 0.82, with a sensitivity of 0.75, specificity of 0.83, and an overall accuracy of 0.81. Similarly, discrimination between controls and patients with mild ataxia showed an AUC of 0.82, with sensitivity of 0.69, specificity of 0.90, and accuracy of 0.82. In contrast, discrimination between mild and severe ataxia showed lower performance, with an AUC of 0.67, sensitivity of 0.88, specificity of 0.45, and accuracy of 0.68.

**Figure 3.**
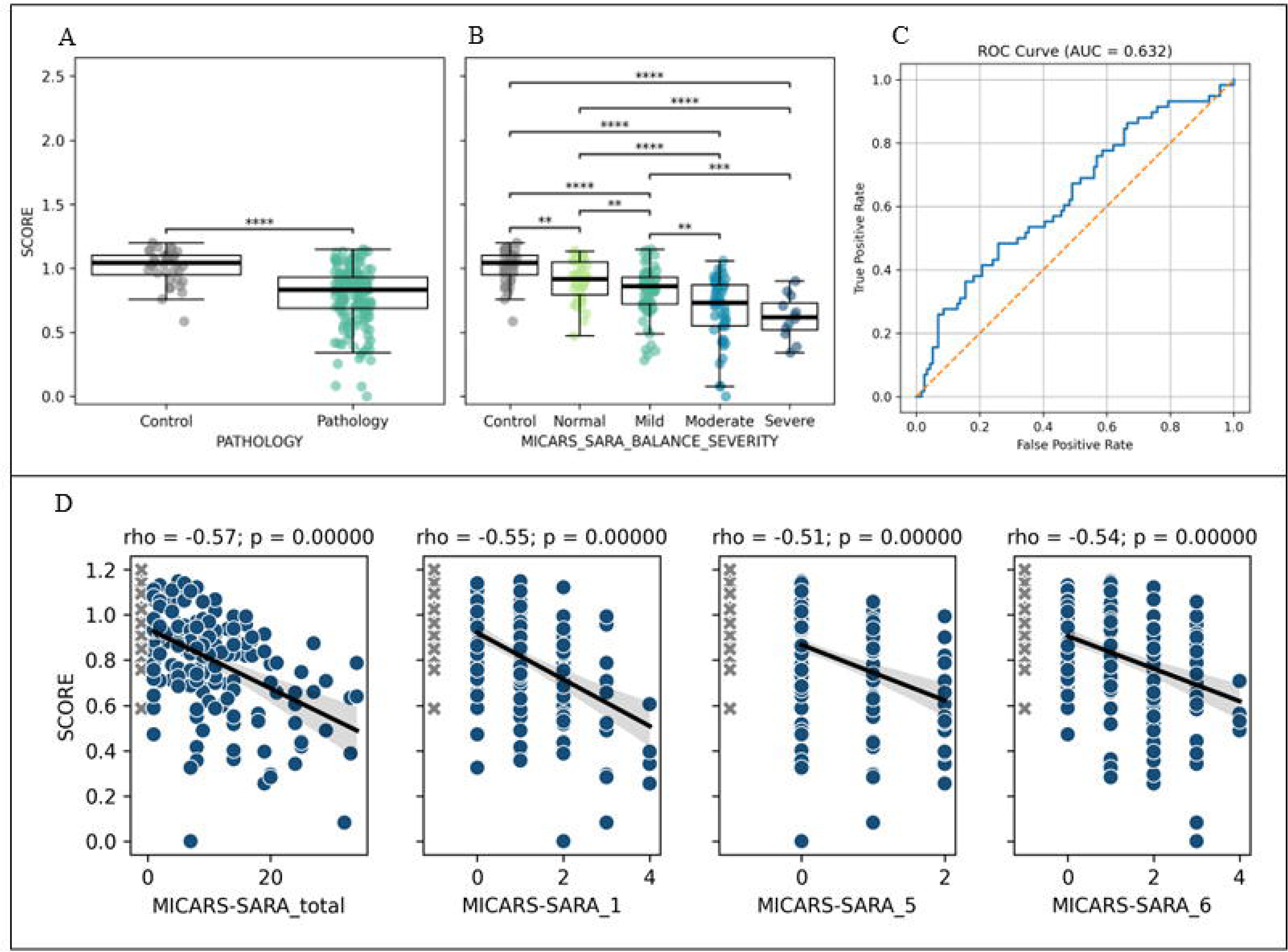
Digital Romberg’s test. (A) Comparison of the normalized digital Romberg’s test between healthy controls and patients with PN. (B) Comparison of the normalized digital Romberg’s test across health controls and patient subgroups stratified by ataxia severity, demonstrating a progressive reduction in the score with increasing clinical severity. Boxes indicate median and interquartile range. Statistical significance is denoted as p< 0.05 (*) p < 0.01 (**), p < 0.001 (***). and p< 0.0001 (****). (C) Receiver operating characteristic (ROC) analysis of the score for discriminating patients with ataxia. The solid blue line represents the model performance while the dashed diagonal line indicates chance level. (D) Correlation of digital Romberg’s test and items of: MICARS and SARA scale. Scatter plots showing Spearman correlations between the instability score and MICARS and SARA total score and selected subitems (1, 5, and *6).* With regression lines and 95% confidence intervals. PN: peripheral neuropathy: MICARS: Modified International Cooperative Ataxia Rating Scale: SARA Scale for the Assessment and Rating of Ataxia

The digital Romberg’s test also demonstrated meaningful clinical validity (Figure 3D), showing moderate negative correlations with MICARS-SARA 5 (ρ = –0.51) and 6 (ρ = –0.54), indicating that greater postural sway and reduced steadiness were associated with lower stability scores. Also, an association was observed with MICARS-SARA item 1, which evaluates gait ataxia (ρ = –0.55), supporting the notion that the proposed balance score reflects both static and dynamic dimensions of ataxic severity. The overall association with the total MICARS-SARA score was more modest (ρ = –0.57), which may be explained by the fact that the scale also assesses upper limb ataxia, which is not necessarily reflected in Romberg’s test performance.

The digital Romberg’s test significantly discriminated between patient groups based on their performance during the Romberg test under both eyes-open and eyes-closed conditions, shown in Supplementary Figure 1A and 1B, with lower scores observed in higher ataxia groups under eyes-closed condition. The digital Romberg’s test also demonstrated significant discrimination among the control, normal, mild and moderate groups (Supplementary Figure 1C). Patients with severe ataxia, defined by MICARS-SARA item 1, were excluded from this last analysis, as there were only two patients assigned to this group, and both were allocated to the training set, leaving no severe cases in the test cohort.

### Correlation analysis between digital Romberg’s test and conventional clinical scales

Supplementary Figure 2 displays the correlations between digital Romberg’s test, clinical scales and PROMS. The digital Romberg’s test showed moderate negative correlations with MICARS and SARA balance-related items, particularly those assessing postural control, while correlations with other clinical and functional scales were generally low. The limited association with global disability measures may reflect the fact that postural instability represents only one of multiple deficits contributing to overall disability and therefore does not fully capture the functional impairment of these patients.

### Digital Romberg’s test differentiates across PN groups

Figure 4 shows the performance of digital Romberg’s test across different PN subtypes. Significant differences were observed between controls and PN patients, with higher values in healthy controls and lower values across all PN subtypes. Among PN, digital Romberg’s test value was significantly lower in CANOMAD and AN subgroups compared to CIDP, MMN and IgM-MGUS, although distributions were partially overlapped, indicating that digital Romberg’s test reflects disease presence and differentiates between pathologies with different ataxia components.

**Figure 4.**
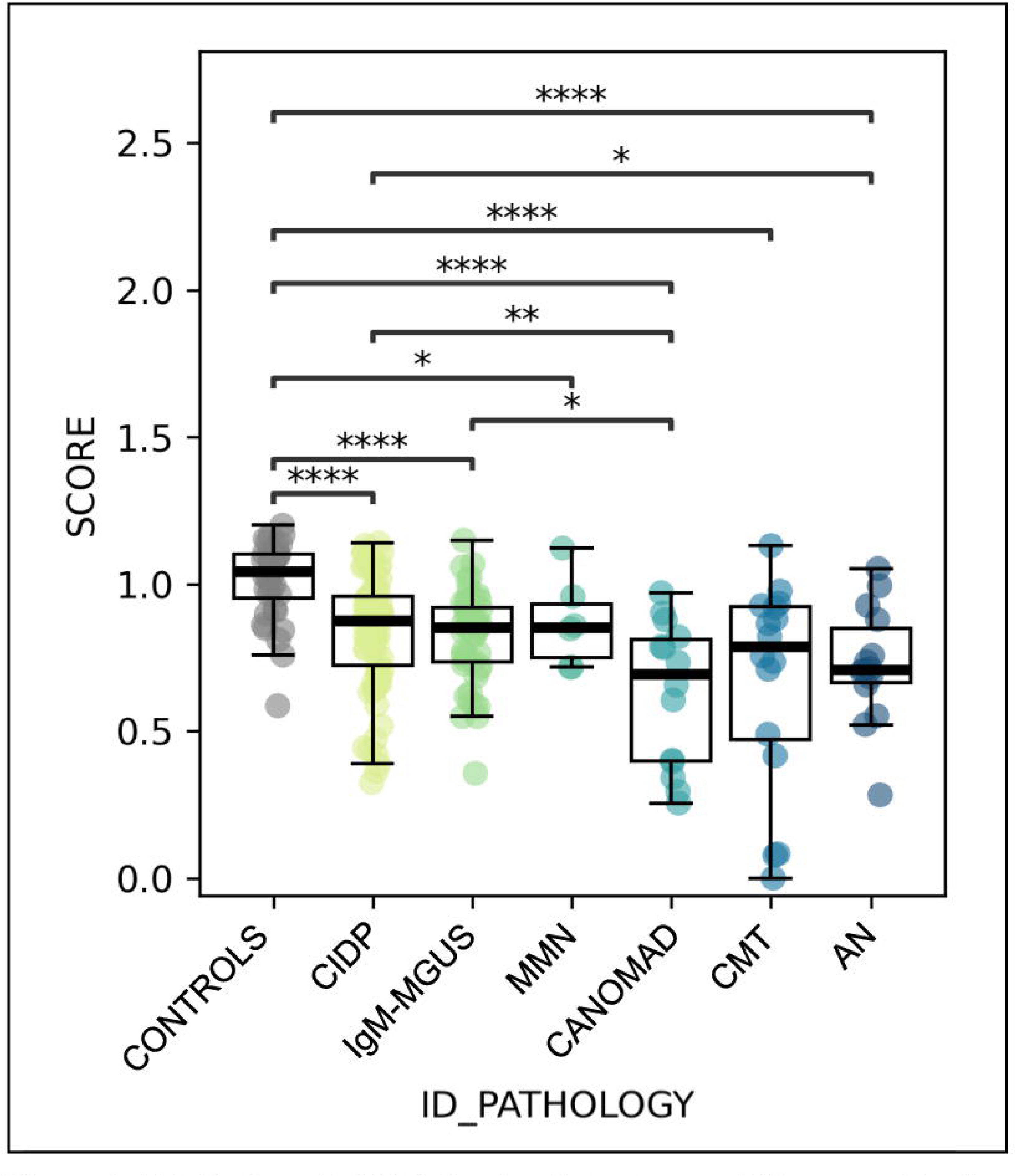
Distribution of digital Romberg’s test across PN groups. Boxplots represents the score values for each pathology group with individual data points overlaid. Points represent individual participants; boxes indicate median and interquartile range. Statistical significance is denoted as p < 0.05 (*), p < 0.01 (**), p < 0.001 (***), and p < 0.0001 (****). PN: peripheral neuropathy; CIDP: chronic inf1ammatory demyelinating polyneuropathy; IgM-MGUS: monoclonal gammopathy patients of undetermined significance associated with IgM; MMN: multifocal motor neuropathy; CANOMAD: chronic ataxic neuropathy, ophthalmoplegia, immunoglobulin M [IgM] paraprotein; CMT: charcot-marie-tooth; AN: autoimmune nodopathy.

### Longitudinal analysis of digital Romberg’s test

To evaluate the clinical relevance of longitudinal changes in the digital Romberg’s test, patients were stratified according to meaningful variations in the score, defined as worsening or improvement if the score decreased or increased in ≥25% of global punctuation (Figure 5). All possible pairs of longitudinal assessments were generated for each patient to identify pairs meeting this threshold, resulting in 53 worsening pairs and 123 improvement pairs. As multiple pairs could originate from the same individual, these corresponded to a total of 10 worsening patients and 25 improvement patients. The patients who worsened according to the digital Romberg’s test showed an increase of +1.42 points (8.9%) (Figure 5A) in MICARS-SARA total score, indicating clinical deterioration. Patients classified as improvers based on their digital score demonstrate a decrease in MICARS-SARA punctuations reflecting their clinical improvement translated into a decrease of −0.21 points (−1.7%) (Figure 5B).

**Figure 5.**
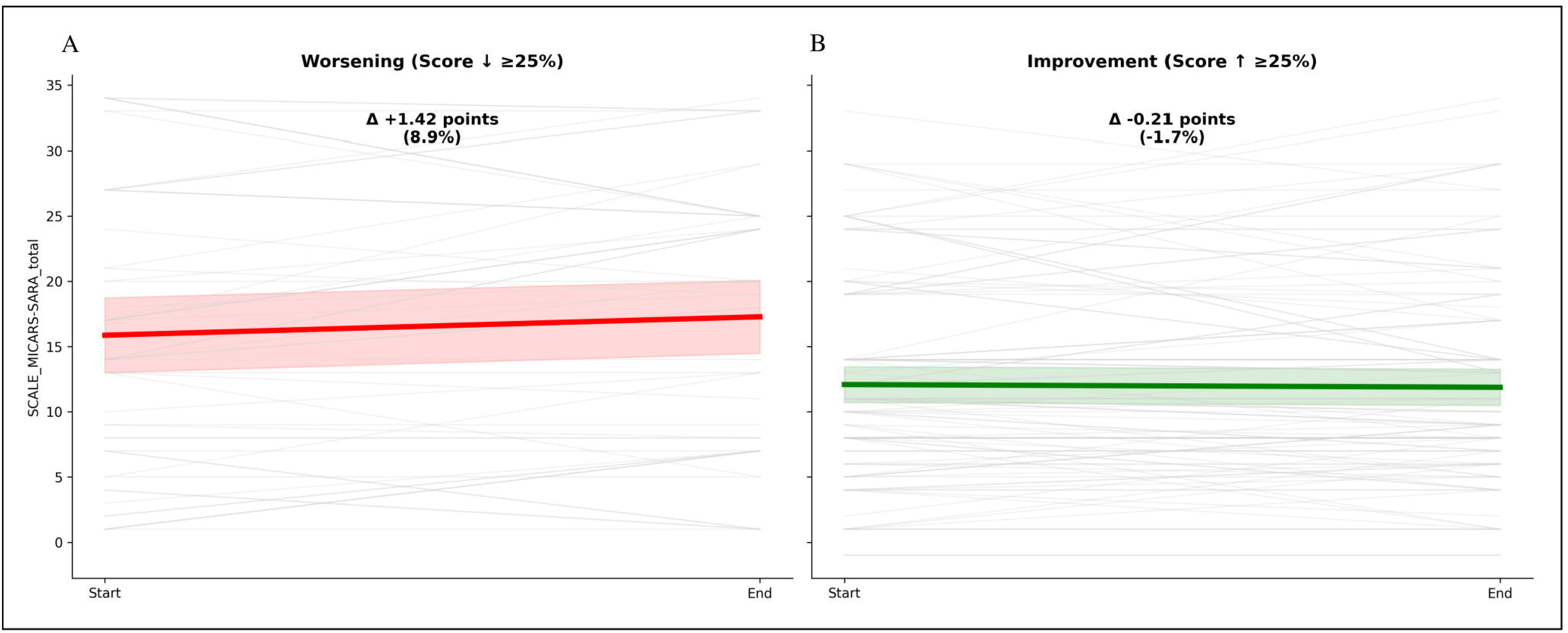
Longitudinal changes in digital Romberg’s test and clinical severity. Changes in MICARS SARA total score according to longitudinal variations in the digital Romberg’s test between visits. Pati ents were stratified based on relative changes in the digital score: worsening (≥25% decrease) and improvement (≥25% increase). Bar plots represent mean changes in MICARS SARA scores, with error bars indicating standard deviation. Positive values indicate clinical worsening, whereas negative values indicate improvement.

To explore how longitudinal changes in the digital Romberg’s test are related to patient clinical status, we stratified assessments into quartiles according to the score and examined the distribution of clinical scales and PROMS (Figure 6). Higher digital Romberg’s test quartiles were associated with less disability and better quality of life, reflected by iRODS and EQ-5D values, and lower impairment, as indicated by reduced MICARS-SARA and INCAT scores. Several pairwise comparisons between quartiles reached statistical significance. MRC values showed minimal variation across digital Romberg’s test quartiles and vigorimeter measurements showed greater variability between quartiles, which correlates with the poor association with these measures as observed in Supplementary Figure 1.

**Figure 6.**
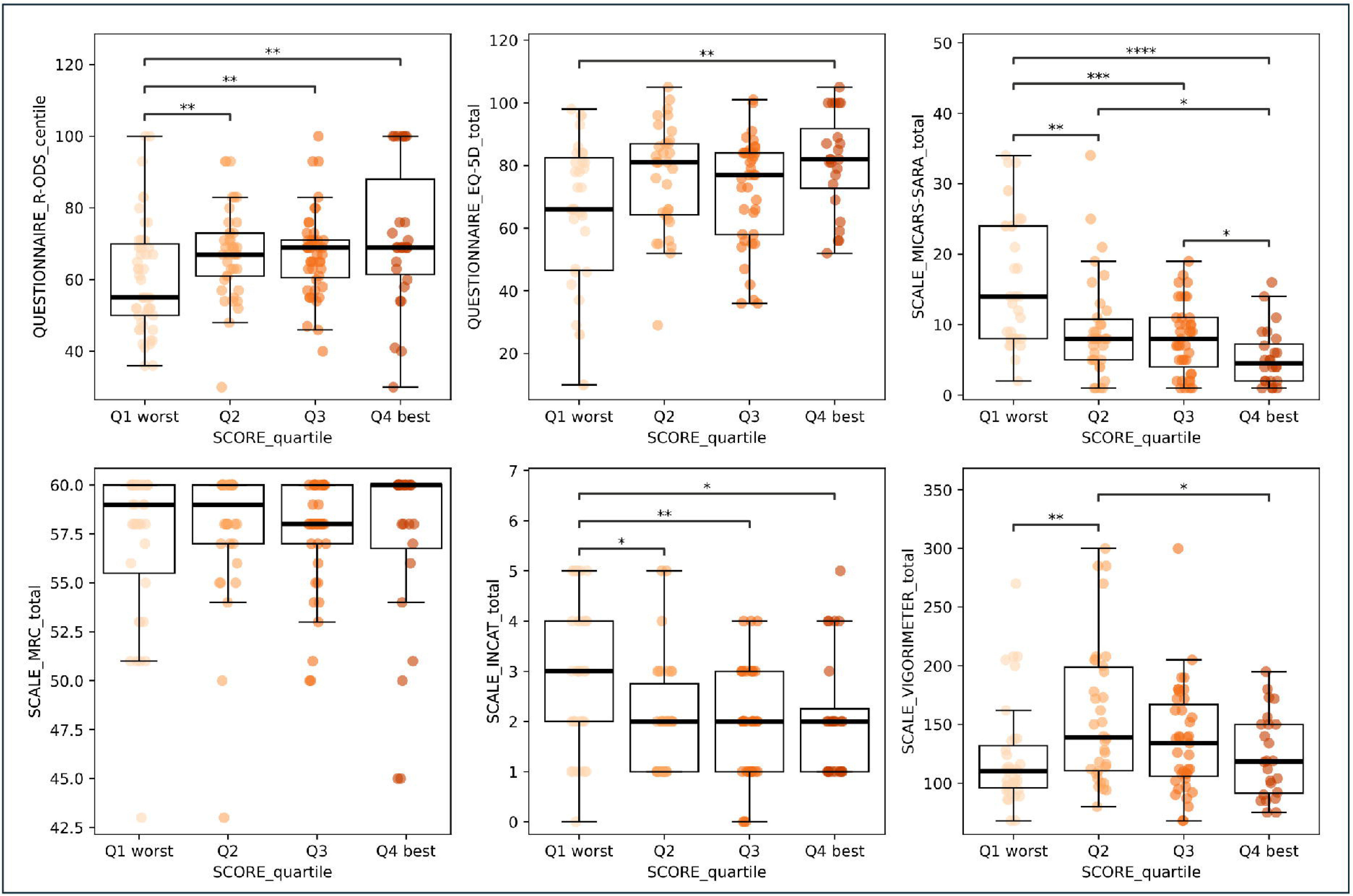
Clinical scales and patient-reported outcomes across quartiles of the digital Romberg’s test score. Boxplots showing the distribution of clinical scales and PROMS across quartiles of the digital Romberg’s test score (Ql: lowest score and worst balance impairment; Q4: highest score and best balance impai1ment. Significant pairwise differences between quartiles are indicated by brackets (p<0.05, p<0.01, p<0.001, ****p<0.0001).

Figure 7 displays the longitudinal trajectories of iRODS centile and digital Romberg’s test over time stratified by change categories: worsened (Δ ≤ −8), stable (|Δ| < 8), improved (Δ ≥ 8). Digital Romberg’s test changes were analyzed within each group, showing trends consistent with the observed clinical changes. In the worsened group, iRODS showed a significant decrease over time, followed by a similar trend in the digital Romberg’s test although not statistically significant. The improved group exhibited a significant increase in iRODS, with a similar trend for the score. The stable group, as expected, did not show significant changes in either measure. Overall, both metrics followed concordant directional changes across subgroups, with iRODS being more sensitive to longitudinal change.

**Figure 7.**
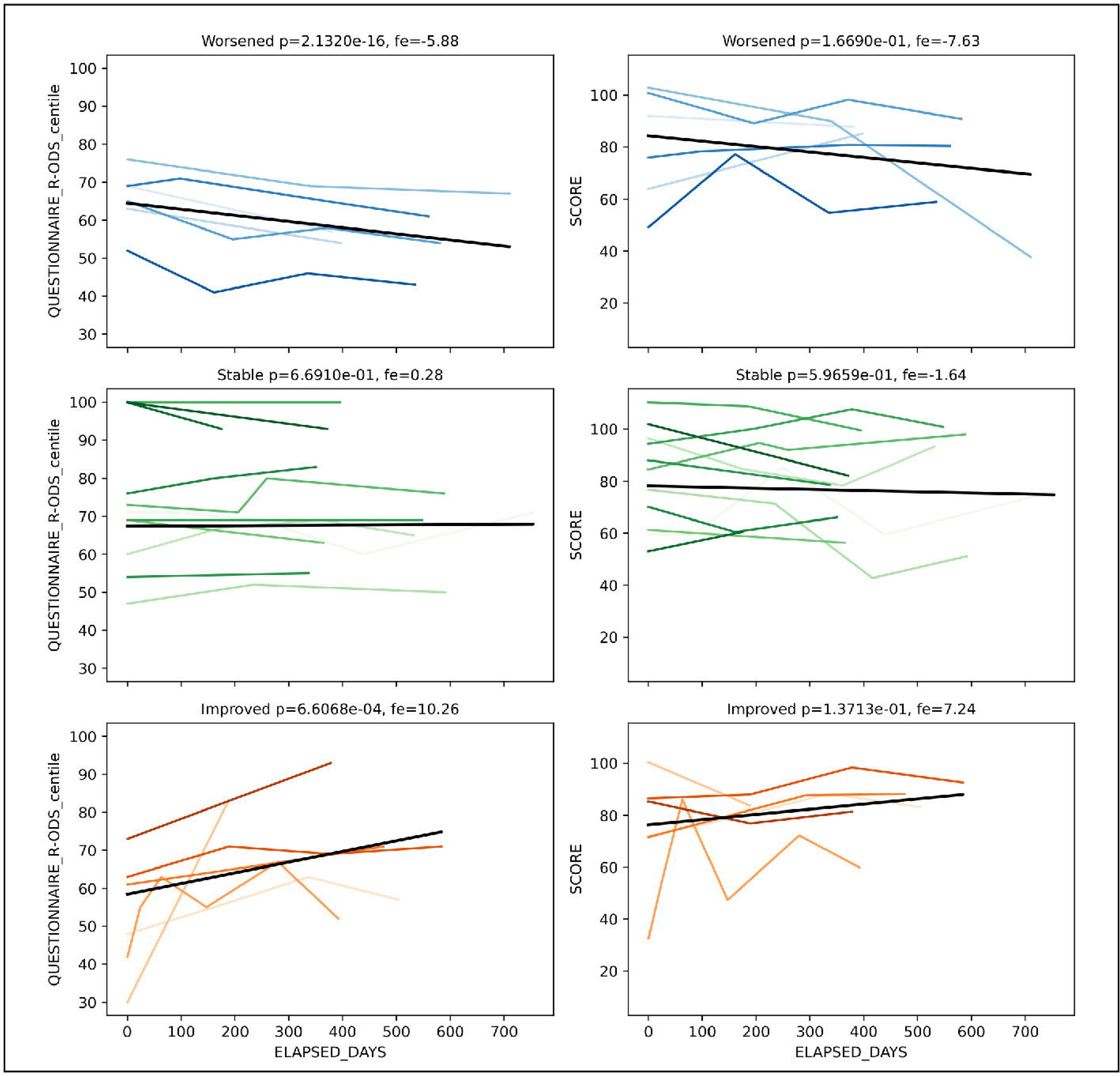
Longitudinal evolution of iRODS centile and digital Romberg’s test according to clinical change categories. Patients were stratified based on the minimal clinically important difference (MCID = 8 points) in R-ODS centile between visits into three groups: Worsened (Δ ≤8), Stable (Δ| < 8), and Improved (Δ ≥ 8). For each subgroup, individual patient trajectories are shown as colored lines, while the black line represents the estimated 1nean trajectory fro1n linear mixed-effects models (LME), including fixed effects of time and rando1n intercepts for patients. The left panels display iRODS centile over time, and the right panels show digital Romberg’s test over time. Reported p-values and fixed effects (fe) correspond to the time effect in each LME model.

## Discussion

Our study shows that digital biomechanical assessment of stance is useful to quantify balance impairment in a large, heterogeneous cohort of patients with PN. Static balance features derived from inertial and plantar pressure sensors and integrated in an AI-generated digital Romberg’s test captured alterations of static posture and correlated with clinical ataxia severity. These findings highlight the potential of this technology to quantify postural instability with precision and reproducibility. Unlike conventional clinical assessments, which often rely on subjective interpretation, the digital Romberg’s test provides an accurate and simple metric that can quantify alterations in postural stability to monitor their progression.

One of the major unmet needs in the clinical management of PN patients is the lack of reliable and objective biomarkers for gait and balance assessment,^31^ which are essential for an accurate diagnosis, prognosis and disease activity monitoring. Current clinical assessments, such as the Romberg’s test or scales for rating ataxia severity are inherently subjective and rely on clinical interpretation, introducing variability and limiting the sensitivity to subtle changes over time. Although timed performance tests, such as two minutes walking test or timed up-and-go tests, provide objective quantitative data, they primarily capture global motor performance and cannot characterize the complete spectrum of biomechanical alterations underlying postural instability. As a result, they may fail to detect compensatory movement patterns or subtle postural control deficits.^32^ This lack of standardization hinders the ability to consistently evaluate disease progression or treatment response, for example, in clinical trials where precise and reproducible outcome measures are essential. The development of objective biomarkers for assessing balance impairment represents a significant step toward filling this gap, which can offer potential to transform routine clinical management by enabling more accurate, quantitative, and reproducible assessments of postural control.

In the context of the current absence of disease-activity biomarkers and relatively poor performance of clinical scales, our main objective was to quantitatively assess balance impairments using a multiparametric wearable system in a cohort of patients with PN. First, we demonstrated that this technology is able to detect significant alterations in trunk sway and center-of-pressure deviation during the Romberg’s test under both open and closed-eyes conditions. We also developed and validated the digital Romberg’s test, an instability score to objectively quantify balance impairment in PN patients, assess its ability to differentiate PN subtypes, evaluate its correlation with conventional clinical scales, and characterize its longitudinal behavior.

Our findings align with prior studies performed in other neurological patients, including patients with diabetic peripheral neuropathy, multiple sclerosis, and other autoimmune neuropathies, in which wearable sensors and force platforms have detected postural control deficits.^15–18^ Although single or grouped biomechanical features with wearable sensors correlate with clinical status of patients and may be longitudinally responsive to clinical changes, it is difficult to select a single biomechanical feature that simplifies disease assessment and monitoring. For this reason, to simplify the analysis and collapse all biomechanical metrics into a single number, we created a composite instability score, the digital Romberg’s test, that discriminated between patients and controls and between different levels of ataxia severity. Our approach offers greater scalability, interpretability and clinical applicability in a heterogeneous cohort of PN patients. To our knowledge, this is the first study evaluating trunk and center-of-pressure movement in a single metric that is fed by a multiparametric wearable system in PN.^11^

The digital Romberg’s test, the instability score we developed, combined multiple static features into a single and interpretable metric that may facilitate clinical monitoring and treatment response evaluation. This composite score demonstrated significant differences across all ataxia severities compared to healthy controls, with more discriminative power between mild and moderate ataxia severity under eyes-closed condition as occurs in routine clinical practice when performing the conventional Romberg’s test. However, discrimination between moderate and severe groups was reduced. It should be noted that patients in the severe group often exhibited difficulties maintaining posture, leading to more constrained movements. This was reflected in the descriptive analysis by a reduction in trunk sway ellipses, likely representing an adaptive strategy to preserve balance and minimize the risk of near-falls during the assessment. In contrast, plantar sway measures appeared to better capture severity-related differences than trunk sway in this subgroup of patients. Moreover, the smaller sample size of this group compared with the mild and moderate groups may have influenced the observed variability. Importantly, the score also showed good ability to identify the presence of ataxia, supporting its potential as a screening tool by effectively detecting balance impairment and distinguishing patients without it.

Correlation analysis of the digital Romberg’s test supported the clinical relevance of the proposed instability score, showing moderate to strong negative correlations with the MICARS-SARA total score and key subitems. These associations were stronger than those observed with other clinical measures, such as MRC or other PROMS, suggesting that the score is particularly sensitive to postural control deficits rather than global functional impairment. This pattern is further reinforced by the quartile analysis. Lower quartiles were consistently associated with greater ataxia severity, increased disability, reduced functional capacity, and poorer quality of life, while higher quartiles reflected better overall clinical status. However, the strength of these associations varied across measures, likely reflecting the weaker correlations observed for scales not directly related to ataxia. Overall, this correlations’ pattern supports its potential role as an objective biomarker aligned with clinical ataxia scales.

The analysis of digital Romberg’s test across PN subtypes further supports the clinical relevance of the score. Lower values were obtained in pathologies with a more prominent sensory ataxic component, such as CANOMAD and autoimmune nodopathies, consistent with their underlying pathophysiology. Disorders like MMN showed significantly higher score values, reflecting their predominant motor phenotype with limited sensory involvement and ataxia. This pattern aligns with its correlations with ataxia-specific clinical scales, reinforcing its specificity for balance-related impairment.

A critical gap in the field is the lack of objective and quantifiable metrics that allow reliable disease monitoring. For this reason, any biomarker should also show responsiveness to longitudinal changes in disease evolution. The longitudinal analysis of digital Romberg’s test also provided evidence of its potential as a monitoring tool. By defining clinically meaningful thresholds of change (≥25%), we demonstrated that variations in the score are associated with changes in the established clinical scales that measure the ataxia severity. However, the magnitude of these changes in the overall MICARS-SARA total score remained relatively modest. This may reflect that balance-related changes, mainly represented by two items of MICARS-SARA scale, are diluted within the broader combined total score. Digital Romberg’s test also followed concordant trajectories with iRODS across clinical change groups defined by MCID thresholds. Although digital Romberg’s test showed consistent longitudinal trends, iRODS demonstrated greater sensitivity to change. This difference reflects the inherent characteristics of each measure. iRODS is a disability scale designed to capture a broad range of functional limitations in daily activities, while digital Romberg’s test is focused on ataxia and balance impairment. These changes, aligned with conventional clinical scales, highlight the score’s potential utility for monitoring disease progression and treatment response. Moreover, the digital Romberg’s test may be particularly sensitive to subtle longitudinal changes in postural control and balance impairment providing a more detailed and objective assessment of balance impairment over time.

The impairment assessment in PN using wearable devices has been performed in a previous study involving patients with diabetic peripheral neuropathy,^33^ where wearable inertial sensors were used to assess postural control across different stance configurations and surface types, providing more sensitive detection of balance impairments and fall risk. The use of different surfaces combined with wearable sensors used in our study would provide a more detailed understanding of the balance impairment in this group of patients.

Despite the promising results, our study has some limitations that should be considered. First, although the overall cohort was diverse and well-characterized, the sample sizes of specific subgroups, such as patients with CANOMAD or CMT, were relatively small, potentially affecting the generalizability. Future studies with larger and stratified samples would be necessary to validate the utility of wearable-based balance assessments. Second, while Romberg’s test provides a standardized and reproducible balance and stability evaluation, it remains a static task and does not capture the multiple neurological deficits that contribute to overall disability in patients with PN. However, our study, in which multiple biomechanical metrics are collapsed into a single composite metric, provides proof-of-principle that similar analyses may also be useful when applied to multiparametric analysis of other neurological assessments, including gait. Finally, postural instability in advanced ataxia may be underrepresented, due to the small number of patients included and the fact that these patients often rely on compensatory strategies and are unable to complete the Romberg’s test adequately.

In conclusion, our study supports the use of wearable sensors as a reliable and more objective method to assess balance impairment in PN. By enabling quantification of postural instability, which correlates well with clinical scales and exhibits consistent patterns over time, the digital Romberg’s test represents a promising, reproducible biomarker for both clinical trials, routine monitoring and treatment response assessment in patients with ataxia. Future research should focus on integrating this tool into treatment evaluation protocols and exploring its predictive value for functional outcomes.

## Acknowledgements

The authors acknowledge the Department of Medicine at the Universitat Auton oma de Barcelona. The authors also thank all our patients for their support and collaboration. Several authors of this publication are members of the European Reference Network for rare neuromuscular diseases (EURO-NMD).

## Ethics

This study was conducted according to a protocol approved by the Ethics Committee of the Hospital de la Santa Creu i Sant Pau (code IIBSP-NMI-2019-107) in accordance with the declaration of Helsinki. All patients provided written informed consent to participate in the study.

## Bibliography

1. Campbell G, Skubic MA. Balance and gait impairment: Sensor-based assessment for patients with peripheral neuropathy. Clin J Oncol Nurs. 2018;22(3):316–325.

2. Oddsson LIE, Bisson T, Cohen HS, et al. The effects of a wearable sensory prosthesis on gait and balance function after 10 weeks of use in persons with peripheral neuropathy and high fall risk - the walk2Wellness trial. Front Aging Neurosci. 2020;12:592751.

3. Zhang Q, Zhou X, Li Y, Yang X, Abbasi QH. Clinical recognition of sensory ataxia and cerebellar ataxia. Front Hum Neurosci. 2021;15:639871.

4. Caronni A, Picardi M, Pintavalle G, et al. Responsiveness to rehabilitation of balance and gait impairment in elderly with peripheral neuropathy. J Biomech. 2019;94:31–38.

5. Dupont L, Defebvre L, Davion JB, Delval A, Tard C. Postural balance and visual dependence in patients with demyelinating neuropathies differ between acquired and hereditary etiologies. Rev Neurol (Paris). 2025;181(1-2):98–105.

6. Mermelstein S, Joffily L, Ellmers TJ, et al. Romberg test: Differentiating vestibular from somatosensory ataxia. Neurol Sci. 2026;47(2):192.

7. Lanska DJ, Goetz CG. Romberg’s sign: development, adoption, and adaptation in the 19th century. Neurology. 2000;55(8):1201–1206.

8. Galán-Mercant A, Cuesta-Vargas AI. Mobile Romberg test assessment (mRomberg). BMC Res Notes. 2014;7(1):640.

9. Anagnostou E, Kouvli M, Karagianni E, et al. Romberg’s test revisited: Changes in classical and advanced sway metrics in patients with pure sensory neuropathy. Neurophysiol Clin. 2024;54(5):102999.

10. Ver MLP, Gum JL, Glassman SD, Carreon LY. Assessment of standing balance in normal versus cervical spondylotic myelopathy patients. N Am Spine Soc J. 2020;3(100023):100023.

11. Tejada-Illa C, Pegueroles J, Claramunt-Molet M, et al. Digital biomechanical assessment of gait in patients with peripheral neuropathies. J Neuroeng Rehabil. 2025;22(1):159.

12. Claramunt-Molet M, Pegueroles J, Pi-Cervera A, et al. Gait analysis reveals new outcome measures for monitoring disease progression in individuals with late-onset Pompe disease. J Neuroeng Rehabil. Published online March 9, 2026. doi:10.1186/s12984-026-01898-8

13. Tsai YH, Li JH, Lin YC, Ting KC, Chan CT. Machine learning-based approaches for tandem Romberg test of vestibular hypofunction assessment using wearable sensors. In: 2024 10th International Conference on Applied System Innovation (ICASI). IEEE; 2024. doi:10.1109/icasi60819.2024.10548001

14. Noamani A, Riahi N, Vette AH, Rouhani H. Clinical static balance assessment: A narrative review of traditional and IMU-based posturography in older adults and individuals with incomplete spinal cord injury. Sensors (Basel). 2023;23(21):8881.

15. Ahmad I, Noohu MM, Verma S, Azharuddin M, Hussain ME. Validity and responsiveness of balance measures using pedalo®-sensomove balance device in patients with diabetic peripheral neuropathy. J Clin Diagn Res. Published online 2019. doi:10.7860/jcdr/2019/40372.12889

16. Meyer BM, Cohen JG, Donahue N, et al. Chest-based wearables and individualized distributions for assessing postural sway in persons with multiple sclerosis. IEEE Trans Neural Syst Rehabil Eng. 2023;31:2132–2139.

17. Sun R, Moon Y, McGinnis RS, et al. Assessment of postural sway in individuals with multiple sclerosis using a novel wearable inertial sensor. Digit Biomark. 2018;2(1):1–10.

18. Michael MR, van Veen R, Wieske L, Merkies ISJ, van Schaik IN, Eftimov F. Validity and responsiveness of balance measurements using posturography in patients with immune-mediated neuropathies. J Peripher Nerv Syst. 2025;30(2):e70031.

19. Findling O, van der Logt R, Nedeltchev K, Achtnichts L, Allum JHJ. A comparison of balance control during stance and gait in patients with inflammatory and non-inflammatory polyneuropathy. PLoS One. 2018;13(2):e0191957.

20. Dinesh K, White N, Baker L, et al. Disease-specific wearable sensor algorithms for profiling activity, gait, and balance in individuals with Charcot-Marie-Tooth disease type 1A. J Peripher Nerv Syst. 2023;28(3):368–381.

21. Schmahmann JD, Gardner R, MacMore J, Vangel MG. Development of a brief ataxia rating scale (BARS) based on a modified form of the ICARS: Brief Ataxia Rating Scale. Mov Disord. 2009;24(12):1820–1828.

22. Subramony SH. SARA--a new clinical scale for the assessment and rating of ataxia. Nat Clin Pract Neurol. 2007;3(3):136–137.

23. Hamadeh T, van Doormaal PTC, Pruppers MHJ, et al. IgM anti-MAG± peripheral neuropathy (IMAGiNe) study protocol: An international, observational, prospective registry of patients with IgM M-protein peripheral neuropathies. J Peripher Nerv Syst. 2023;28(2):269–275.

24. Ephion Health. https://www.ephion.health/

25. Movesense. https://www.movesense.com/

26. Moticon. https://moticon.com/

27. Gołąb A. Describing center of pressure movement in stabilometry by ellipse area approximation. Physiol Rep. 2022;10(17):e15390.

28. Germanotta M, Mileti I, Conforti I, Del Prete Z, Aprile I, Palermo E. Estimation of human center of mass position through the inertial sensors-based methods in postural tasks: An accuracy evaluation. Sensors (Basel). 2021;21(2):601.

29. Quijoux F, Nicolaï A, Chairi I, et al. A review of center of pressure (COP) variables to quantify standing balance in elderly people: Algorithms and open-access code. Physiol Rep. 2021;9(22):e15067.

30. van Veen R, Wieske L, Lucke I, et al. Assessing deterioration using impairment and functional outcome measures in chronic inflammatory demyelinating polyneuropathy: A post-hoc analysis of the immunoglobulin overtreatment in CIDP trial. J Peripher Nerv Syst. 2022;27(2):144–158.

31. Allen JA, Eftimov F, Querol L. Outcome measures and biomarkers in chronic inflammatory demyelinating polyradiculoneuropathy: from research to clinical practice. Expert Rev Neurother. 2021;21(7):805–816.

32. Iliescu B, Herascu A, Gaita L, Avram VF, Timar B. Associations between diabetic neuropathy and balance impairments in patients with type 2 diabetes: A cross-sectional study. J Clin Med. 2025;14(23):8323.

33. Brognara L, Sempere-Bigorra M, Mazzotti A, Artioli E, Julián-Rochina I, Cauli O. Wearable sensors-based postural analysis and fall risk assessment among patients with diabetic foot neuropathy. J Tissue Viability. 2023;32(4):516–526.

