## Supplementary tables for "Artificial intelligence-generated digital Romberg test for peripheral neuropathy monitoring"

| | Ataxia MICARS-SARA Item 1 (walking capacities) | Definition |
|---|---|---|
| **Normal** | Value 0 | (0) Normal |
| **Mild** | Value 1-2 | (1) Almost normal naturally, but unable to walk with feet in tandem position. (2) Walking without support, but clearly abnormal and irregular. |
| **Moderate** | Value 3-4 | (3) Walking without support but with considerable staggering; difficulties in half turn. (4) Walking with autonomous support no longer possible; the patient uses the episodic support of the wall for 10-meter test. |
| **Severe** | Value ≥ 5 | (5) Walking only possible with one stick. (6)Walking only possible with two special sticks or with a stroller. (7) Walking only with accompanying person. (8) Walking impossible, even with accompanying person (wheelchair). |

**Supplementary Table 1. Ataxia severity classification.** Categorization performed following the score obtained in item 1 of MICARS and SARA scales for ataxia patients and its definition according to the score obtained. MICARS: Modified Internacional Cooperative Ataxia Rating Scale; SARA: Scale for the Assessment and Rating of Ataxia.

| | MICARS 5 (open eyes Romberg) | MICARS 6 (closed eyes Romberg) | Definition |
|---|---|---|---|
| **Normal** | Value 0 | Value 0 | Normal (<10 cm) |
| **Mild** | Value 1 | Value 1 | Oscillations |
| **Moderate** | Value 2 | Value 2 | Moderate oscillations (<10 cm at the level of head) |
| **Severe** | Value 3 | Value 3 | Severe oscillations (>10 cm at the level of head), threatening the upright position |
| **Falling** | Value 4 | Value 4 | Immediate falling |

**Supplementary Table 2. Romberg's test cohorts' classification.** Categorization performed following the score obtained in item 5 and 6 of MICARS scale for ataxia patients and its definition according to the score obtained. MICARS: Modified Internacional Cooperative Ataxia Rating Scale.

| Biomechanical feature | Definition |
|---|---|
| MINOR-AXIS | Minor axis of the ellipse (m) |
| MAJOR-AXIS | Major axis of the ellipse (m) |
| RATIO-XY | Ratio between the minor and major axis of the ellipse |
| AREA-XY | Ellipse area (95% of the points) |
| ELLIPSE-CENTER-X | Ellipse center position, interpreted as the mean position of all points on the x-axis |
| ELLIPSE-CENTER-Y | Ellipse center position, interpreted as the mean position of all points on the y-axis |
| PROM-DISTANCE-XY | Average distance: Path_length/len(data) → average distance between two points |
| VAR-DISTANCE-XY | Variance of the distance between each pair of points |
| PATH-LENGTH-XY | Path traveled by the COP (cumulative point-to-point sum of the distance) |
| RMS-X | RMS of the data on the X-axis |
| RMS-Y | RMS of the data on the X-axis |
| PROM-VEL-XY | Derivative of position → gives velocity, and its mean is calculated |
| PROM-ACC-XY | Derivative of velocity → gives acceleration, and its mean is calculated |
| COM_X, COM_Y | Calculation of the lateral and anteroposterior trunk position signal through double integration of acceleration |
| TRUNK_ACC_ROM | Range of motion of acceleration on the three axes |
| TURNK_ACC_STD | STD (standard deviation) of acceleration on the three axes |
| TRUNK_ACC_VAR | Variance of acceleration on the three axes |
| TRUNK_SWAY-AREA-XY | Area occupied by trunk sway without reducing to 95% |
| TRUNK_SWAY-VELOCITY-XY | Velocity calculated based on total COM path length and signal time |
| TRUNK_PATH-LENGTH-XY | Total path length of the COM |
| TRUNK_MEDIOLATERAL_X | Mediolateral movement amplitude |
| TRUNK_RMS_X | RMS of the COM in X |
| TRUNK_RMS_Y | RMS of the COM in Y |
| TRUNK_ANTERIOPOSTERIOR-SWAY-X | Anteroposterior movement amplitude |
| TRUNK_DIRECTIONAL_X | Median of the position signal in X |
| TRUNK_DIRECTIONAL_Y | Median of the position signal in Y |

**Supplementary Table 3. Definition of static features measured by biomechanical sensors during the postural control assessment.** The table summarizes the extracted features derived from sensor signals, including measures related to body sway, stability, and postural control, together with a brief description of their biomechanical meaning and units where applicable.

| | | | | | | | | | |
|---|---|---|---|---|---|---|---|---|---|
| | | | | **Ataxia group** | | | | | |
| | | *Control* | *Normal* | *Mild* | *Moderate* | *Severe* | *Falling* | *Not specified* | *p-value* |
| **n (tests)** | | 54 | 112 | 145 | 19 | 6 | - | | |
| **n (patients)** | | 54 | 48 | 5 | 10 | 3 | - | 11 | |
| **Age (years), mean (SD)** | | 59.8 (9.9) | 58.8 (11.9) | 62.8 (13.1) | 60.6 (9.0) | 59.5 (6.8) | - | | 0.096 |
| **Sex, n (%)** | | 18 (33.3) | 70 (61.4) | 95 (65.5) | 7 (36.8) | 6 (100) | - | | <0.001 |
| | | | | Open-eyes group | | | | | |
| **n (tests)** | | 54 | 184 | 67 | 26 | 2 | 3 | | |
| **n (patients)** | | 54 | 75 | 41 | 12 | 2 | 2 | | <0.001 |
| **Age (years), mean (SD)** | | 59.8 (9.9) | 60.5 (13.1) | 63.1 (12.4) | 58.7 (6.7) | 63.5 (0.7) | 61.7 (2.5) | | 0.565 |
| **Sex, n (%)** | Male | 18 (33.3) | 116 (62.4) | 43 (64.2) | 15 (57.7) | 1 (50) | 3 (100.0) | | 0.003 |
| | | | | Closed-eyes group | | | | | |
| **n** | | 54 | 78 | 82 | 61 | 46 | 15 | | |
| **n (patients)** | | 54 | 41 | 5 0 | 43 | 28 | 8 | | <0.001 |
| **Age (years), mean (SD)** | | 59.8 (9.9) | 57.5 (12.2) | 60.9 (14.5) | 64.7 (11.6) | 62.1 (9.3) | 61.0 (5.3) | | 0.020 |
| **Sex, n (%)** | Male | 18 (33.3) | 50 (64.1) | 54 (64.3) | 34 (55.7) | 31 (67.4) | 9 (60.0 | | 0.003 |

**Supplementary Table 4. Demographic characteristics of patients included in the study.** Data are presented according to the score obtained in item 1 of MICARS-SARA scale and according to testing condition (open eyes vs. closed eyes). Continuous variables are expressed as mean ± standard deviation (SD) and categorical variables are presented as counts and percentages (%). MICARS-SARA: Modified International Cooperative Ataxia Rating Scale - Scale for the Assessment and Rating of Ataxia.

| Feature | β Path Open | p Path Open | p Path Open Corrected | β Path Close | p Path Close | p Path Close Corrected |
|---|---|---|---|---|---|---|
| **INSOLE_COP_AREA-XY_L** | 0,0010 | 0,0185 | **0,0467** | 0,0020 | 0,0001 | **0,0012** |
| **INSOLE_COP_AREA-XY_R** | 0,0007 | 0,1019 | 0,1882 | 0,0019 | 0,0297 | 0,0606 |
| **INSOLE_COP_CORR-OSCILLATION-XY** | - 0,0629 | 0,1523 | 0,2707 | - 0,0142 | 0,7731 | 0,8689 |
| **INSOLE_COP_ELLIPSE-CENTER-X_L** | - 0,0182 | 0,2101 | 0,3056 | - 0,0027 | 0,8630 | 0,8817 |
| **INSOLE_COP_ELLIPSE-CENTER-X_R** | - 0,0249 | 0,0842 | 0,1617 | - 0,0096 | 0,5150 | 0,6206 |
| **INSOLE_COP_ELLIPSE-CENTER-Y_L** | 0,0113 | 0,2173 | 0,3068 | 0,0078 | 0,4099 | 0,5351 |
| **INSOLE_COP_ELLIPSE-CENTER-Y_R** | 0,0027 | 0,7508 | 0,8190 | 0,0046 | 0,5906 | 0,6940 |
| **INSOLE_COP_MAJOR-AXIS-XY_L** | 0,0023 | 0,0505 | 0,1155 | 0,0052 | 0,0001 | **0,0012** |
| **INSOLE_COP_MAJOR-AXIS-XY_R** | 0,0012 | 0,3304 | 0,4173 | 0,0037 | 0,0506 | 0,0951 |
| **INSOLE_COP_MINOR-AXIS-XY_L** | 0,0157 | 0,0001 | **0,0020** | 0,0280 | <0,0001 | **<0,0001** |
| **INSOLE_COP_MINOR-AXIS-XY_R** | 0,0115 | 0,0028 | **0,0084** | 0,0221 | <0,0001 | **0,0001** |
| **INSOLE_COP_PROM-ACC-XY_L** | 0,0151 | 0,4352 | 0,5356 | 0,0272 | 0,2741 | 0,3790 |
| **INSOLE_COP_PROM-ACC-XY_R** | - 0,0242 | 0,1639 | 0,2810 | 0,0882 | 0,0009 | **0,0035** |
| **INSOLE_COP_PROM-DISTANCE-XY_L** | 0,0001 | 0,7406 | 0,8190 | 0,0004 | 0,2680 | 0,3790 |
| **INSOLE_COP_PROM-** | - 0,0028 | 0,0000 | **0,0002** | - 0,0019 | 0,0014 | **0,0042** |

| | | | | | | |
|---|---|---|---|---|---|---|
| **DISTANCE-XY_R** | | | | | | |
| **INSOLE_COP_PROM-VEL-XY_L** | 0,0112 | 0,7406 | 0,8190 | 0,0436 | 0,2680 | 0,3790 |
| **INSOLE_COP_PROM-VEL-XY_R** | - 0,2815 | 0,0000 | **0,0002** | - 0,1923 | 0,0014 | **0,0042** |
| **INSOLE_COP_RATIO-XY_L** | - 0,1124 | 0,0002 | **0,0025** | - 0,1015 | 0,0004 | **0,0023** |
| **INSOLE_COP_RATIO-XY_R** | - 0,0925 | 0,0016 | **0,0068** | - 0,1169 | <0,0001 | **0,0003** |
| **INSOLE_COP_RMS-X_L** | 0,0016 | 0,8313 | 0,8532 | 0,0018 | 0,8134 | 0,8689 |
| **INSOLE_COP_RMS-X_R** | 0,0132 | 0,0733 | 0,1530 | 0,0185 | 0,0117 | **0,0262** |
| **INSOLE_COP_RMS-Y_L** | 0,0011 | 0,8829 | 0,8829 | 0,0016 | 0,8384 | 0,8757 |
| **INSOLE_COP_RMS-Y_R** | 0,0143 | 0,0536 | 0,1169 | 0,0180 | 0,0129 | **0,0275** |
| **TRUNK_ACC_STDX** | 0,0454 | 0,0010 | **0,0046** | 0,0509 | 0,0037 | **0,0103** |
| **TRUNK_ACC_STDY** | 0,0807 | 0,0004 | **0,0035** | 0,0828 | 0,0011 | **0,0038** |
| **TRUNK_ACC_STDZ** | 0,0481 | 0,0020 | **0,0072** | 0,0691 | 0,0002 | **0,0012** |
| **TRUNK_ANTERIOPOSTERIOR-SWAY-Y** | 0,0483 | 0,0051 | **0,0144** | 0,0484 | 0,0044 | **0,0109** |
| **TRUNK_AREA-XY** | 0,0008 | 0,1878 | 0,2828 | 0,0011 | 0,0583 | 0,1016 |
| **TRUNK_DIRECTIONAL-X** | <0,0001 | 0,1885 | 0,2828 | <0,0001 | 0,8014 | 0,8689 |
| **TRUNK_DIRECTIONAL-Y** | <0,0001 | 0,8354 | 0,8532 | <0,0001 | 0,2144 | 0,3251 |
| **TRUNK_ELLIPSE-CENTER-X** | <0,0001 | 0,1885 | 0,2828 | <0,0001 | 0,8014 | 0,8689 |
| **TRUNK_ELLIPSE-CENTER-Y** | 0,0000 | 0,8354 | 0,8532 | - 0,0000 | 0,2144 | 0,3251 |
| **TRUNK_MAJOR-AXIS-XY** | 0,0030 | 0,0107 | **0,0286** | 0,0040 | 0,0041 | **0,0107** |
| **TRUNK_MEDIOLATERAL-X** | 0,0255 | 0,0021 | **0,0073** | 0,0342 | 0,0006 | **0,0028** |

| | | | | | | |
|---|---|---|---|---|---|---|
| **TRUNK_MINOR-AXIS-XY** | 0,0145 | 0,0009 | **0,0046** | 0,0151 | 0,0011 | **0,0038** |
| **TRUNK_PATH-LENGTH-XY** | 0,1411 | 0,0007 | **0,0044** | 0,1562 | 0,0005 | **0,0024** |
| **TRUNK_RATIO-XY** | 0,0139 | 0,6208 | 0,7268 | 0,0033 | 0,9093 | 0,9093 |
| **TRUNK_RMS-X** | 0,0064 | 0,0017 | **0,0068** | 0,0096 | 0,0002 | **0,0017** |
| **TRUNK_RMS-Y** | 0,0127 | 0,0028 | **0,0084** | 0,0116 | 0,0080 | **0,0189** |
| **TRUNK_SWAY-AREA-XY** | 0,0008 | 0,1878 | 0,2828 | 0,0011 | 0,0583 | 0,1016 |
| **TRUNK_SWAY-VELOCITY-XY** | 0,0353 | 0,0007 | **0,0044** | 0,0386 | 0,0005 | **0,0025** |
| **TRUNK_CORR-OSCILLATION-XY** | - 0,0427 | 0,0765 | 0,1531 | - 0,0159 | 0,5049 | 0,6206 |
| **INSOLE_COP_VAR-DISTANCE-XY_L** | <0,0001 | 0,3234 | 0,4173 | <0,0001 | 0,1062 | 0,1722 |
| **INSOLE_COP_VAR-DISTANCE-XY_R** | <0,0001 | 0,5721 | 0,6865 | <0,0001 | 0,4904 | 0,6206 |
| **TRUNK_ACC_VARX** | 0,0131 | 0,2701 | 0,3704 | 0,0193 | 0,2936 | 0,3943 |
| **TRUNK_ACC_VARY** | 0,0382 | **0,0476** | 0,1141 | 0,0482 | 0,0354 | 0,0694 |
| **TRUNK_ACC_VARZ** | 0,0170 | 0,3229 | 0,4173 | 0,0272 | 0,0680 | 0,1142 |

**Supplementary Table 5. Association between wearable-derived balance features and patient status (PN vs controls).** Linear mixed-effects model results evaluating the association between wearable-derived balance features and patient status during the Romberg's test under eyes-open and eyes-closed conditions. In green significant features p-values related to COP features, in orange significant p-values related to trunk sway features. A significance threshold of $p < 0.05$ was applied to identify features showing statistically significant differences between patient groups and controls.

| Feature | β Path Open | p Path Open | p Path Open Corrected | β Path Close | p Path Close | p Path Close Corrected |
|---|---|---|---|---|---|---|
| INSOLE_COP_AREA-XY_L | 0,0455 | 0,3131 | 0,3131 | 0,2656 | 0,0057 | **0,0087** |
| INSOLE_COP_MINOR-AXIS-XY_L | 0,2535 | **0,0120** | 0,0208 | 0,3964 | 0,0040 | **0,0075** |
| INSOLE_COP_MINOR-AXIS-XY_R | 0,2775 | **0,0124** | 0,0208 | 0,7141 | 0,0006 | **0,0045** |
| INSOLE_COP_PROM-DISTANCE-XY_R | 0,6255 | **0,0002** | 0,0024 | 0,5394 | 0,0012 | **0,0051** |
| INSOLE_COP_PROM-VEL-XY_R | 0,6255 | **0,0002** | 0,0024 | 0,5394 | 0,0012 | **0,0051** |
| INSOLE_COP_RATIO-XY_L | 0,4663 | **0,0016** | 0,0076 | 0,4023 | 0,0059 | **0,0087** |
| INSOLE_COP_RATIO-XY_R | 0,2583 | **0,0290** | 0,0369 | 0,3785 | 0,0066 | **0,0091** |
| TRUNK_ACC_STD X | 0,2097 | **0,0142** | 0,0208 | 0,3439 | 0,0077 | **0,0100** |
| TRUNK_ACC_STD Y | 0,4607 | **0,0006** | 0,0035 | 0,7885 | 0,0001 | **0,0026** |
| TRUNK_ACC_STD Z | 0,2068 | **0,0133** | 0,0208 | 0,2054 | 0,0280 | **0,0342** |
| TRUNK_ANTERIOPOSTERIOR-SWAY-Y | 0,1838 | **0,0291** | 0,0369 | 0,3229 | 0,0054 | **0,0087** |
| TRUNK_MAJOR-AXIS-XY | 0,1193 | 0,0764 | 0,0854 | 0,1621 | 0,0335 | **0,0388** |
| TRUNK_MEDIOLATERAL-X | 0,1580 | **0,0415** | 0,0493 | 0,1610 | 0,0675 | 0,0718 |
| TRUNK_MINOR-AXIS-XY | 0,2476 | **0,0092** | 0,0208 | 0,3750 | 0,0030 | **0,0075** |
| TRUNK_PATH-LENGTH-XY | 0,2386 | **0,0100** | 0,0208 | 0,3948 | 0,0023 | **0,0072** |
| TRUNK_RMS-X | 0,1117 | 0,0936 | 0,0988 | 0,1511 | 0,0685 | 0,0718 |
| TRUNK_RMS-Y | 0,2944 | **0,0050** | 0,0189 | 0,3416 | 0,0038 | **0,0075** |
| TRUNK_SWAY-VELOCITY-XY | 0,2381 | **0,0100** | 0,0208 | 0,3947 | 0,0023 | **0,0072** |

**Supplementary Table 6. Association between wearable-derived balance features and ataxia severity.** Linear mixed-effects model results evaluating the association between wearable-derived balance features and ataxia severity during the Romberg's test under eyes-open and eyes-closed conditions. In green significant features p-values related to COP features, in orange significant p-values related to trunk sway features. A significance threshold of $p < 0.05$ was applied to identify features showing statistically significant differences between patient groups.
