## Supplementary figures and images for "Artificial intelligence-generated digital Romberg test for peripheral neuropathy monitoring"

### Supplementary Figure 1

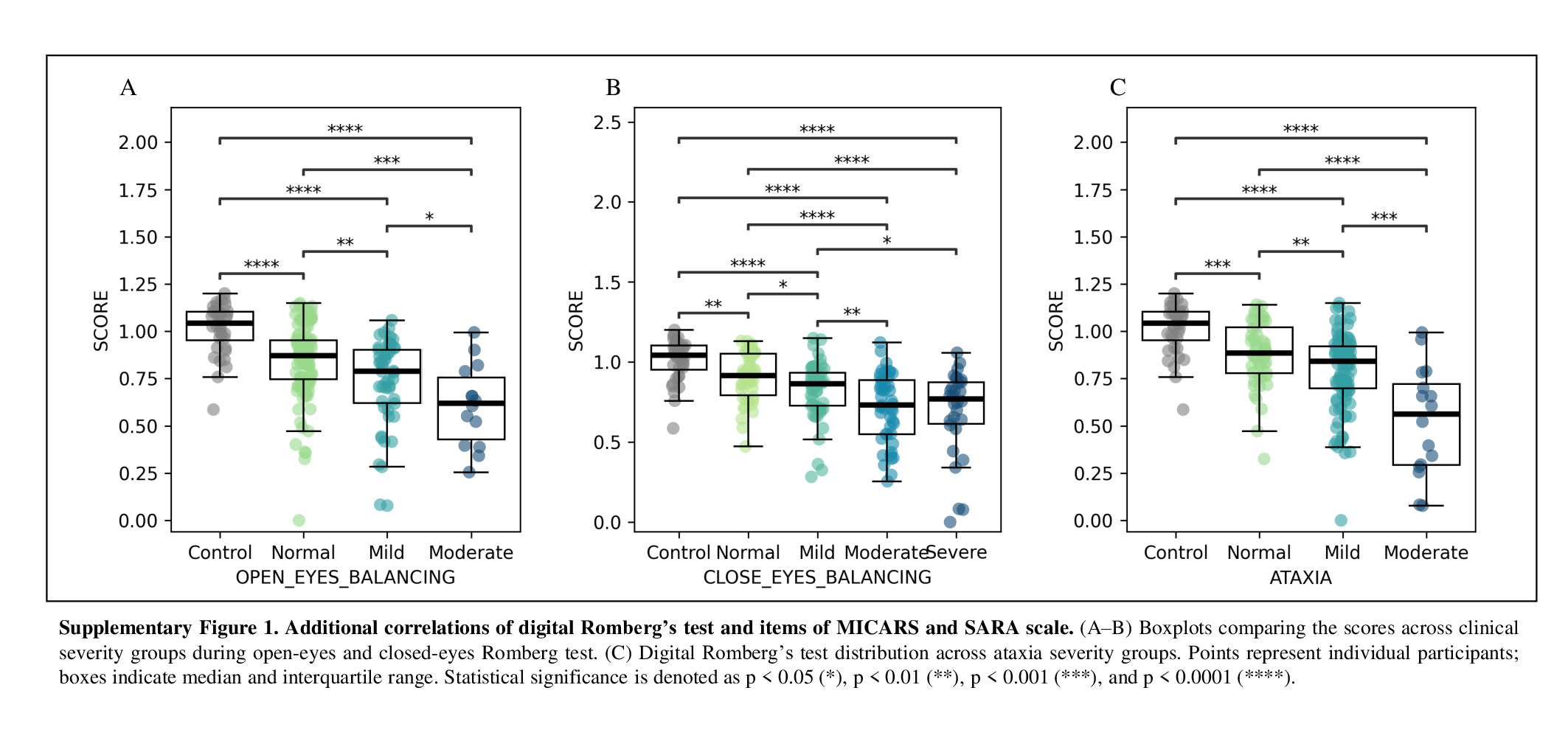

### Supplementary Figure 2

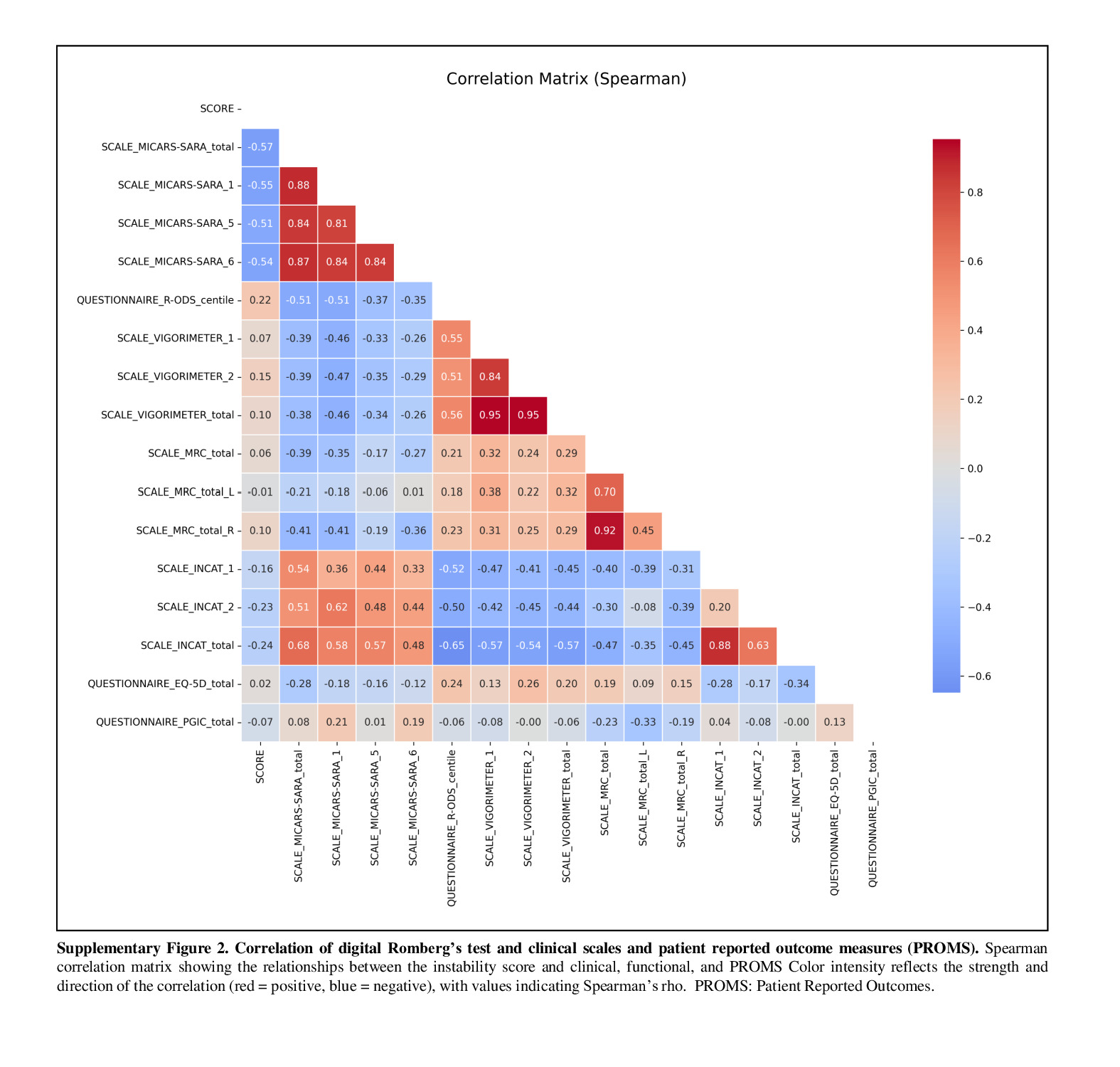
